# Efficient Triage and Improved Patient Care: Five years of Teledermatology in Styria

**DOI:** 10.1101/2025.07.22.25331267

**Authors:** Arzberger Edith, Bordag Natalie, Hofmann-Wellenhof Elena, Freytag Leo, Hofmann-Wellenhof Rainer

## Abstract

**Introduction:** To improve dermatological care in underserved rural regions, the “Teledermatology in Styria” project launched on January 1^st^, 2020. This project digitally connected general practitioners (GP) with dermatologists (DERM), funded by Styrian Health Fund and supported by a partnership between Styrian Medical Association, Austrian Health Insurance Fund Styria, Universities’ Department of Dermatology, and e-derm-consult GesmbH.

**Methodology:** The project allows GPs to submit clinical and dermoscopic images of patient skin conditions, with relevant clinical information, via a “store-and-forward” teledermatology service to DERM, who responds with diagnosis and treatment recommendations. After five years, the collected data was analyzed, and acceptance among patients and doctors was assessed using questionnaires.

**Results:** Of 5119 cases, 19% required no therapy, while 61% were managed by their GPs following teledermatological consultation. Referrals included 12% for routine dermatological appointments, 3% for urgent appointments, and 2% for hospital visits. The case spectrum covered all dermato-venereological conditions, with neoplasms accounting for only 34%. Patient satisfaction exceeded 95%, based on 692 returned questionnaires.

**Conclusion:** Only 17% of patients required additional specialist dermatological examination after teledermatology. The notably faster diagnosis and the surprisingly high level of patient satisfaction highlight the benefits of teledermatological consultations.

## Introduction

Even with Austria’s strong medical infrastructure, widespread dermatological care, particularly in rural regions, is characterized by shortages, resulting in several month long waiting periods for dermatologist (DERM) appointments.

Dermatology and venereology became elective subjects in the Austrian general medicine training ordinancen^1^. Yet, practical experience shows that 10-20% of all general practitioners (GPs) patients present with a dermatological condition^2–4^. Inflammatory conditions are most common, though skin tumours prevalence is increasing^5,6^, due to changing lifestyles and demographics^7,8^. Early and rapid skin cancer detection is possible with dermatoscopy^9,10^. Timely intervention reduces sick leave, avoids costly advanced therapies, and significantly improves affected individuals’ quality of life^11,12^.

Teledermatology is emerging globally as an indispensable, fully integrated component of dermatological care^13^, particularly given the skin’s inherent accessibility and dermatology being a morphological discipline. Both clinical experience and literature confirm that teledermatological diagnosis is non-inferior to in-person assessment^14–16^.

Teledermatology links DERM digitally with GPs or patients via: (a) asynchronous store-and-forward (SF), (b) synchronous real-time-applications (RA) or (c) in hybrid models^17,18^. Each approach offers distinct benefits and disadvantages. SF offers low resource use, straightforward technical needs, and time-independent operation, while RA offers enhanced flexibility during anamnesis and direct discussion of diagnosis and treatment options. The connection of DERM with GPs ensures clinically professional and fast case creation, including medical history and shared therapy management, whereas direct DERM-to-patient links reduce the GPs workload.

A 2018 survey involving 243 Austrian dermatologists (47% male, 55% female) revealed that roughly half had teledermatology experience. Among these, half were for second opinions, and only one-third for initial visits. A substantial 73% considered teledermatology capable of reducing their routine workload^19^.

The “Teledermatology in Styria” project was launched to improve dermatological patient care in rural areas, drawing on experiences from Germany^20^. Initially, the district of Liezen was selected due to its low population density and significant distance from urban centers. Subsequently, the project expanded to include a district with a similar population size but close proximity to the urban center of Graz.

## Methods

### Project progression

The project commenced on January 1^st^, 2020, following approval by the Ethics Committee of the Medical University of Graz (EK 32-228 ex 19/29) and was funded by the Styrian Health Fund. The Styrian Medical Association acted as the project applicant, and the Austrian Health Insurance Fund (ÖGK) reimbursed the medical services. Scientific guidance was provided by the Medical University of Graz and technical implementation by e-derm-consult GesmbH.

GPs underwent 15-30 minute, mostly in-person, training sessions on proper image and case submission via the teledermatology system (security detailed in Appendix 1). This enabled teledermatological requests as alternative to conventional referral for skin conditions. All GPs were provided with a tablet (iPad mini 5, Apple Inc. Cupertino, CA, USA) and a portable, maintenance-free dermoscope with polarization (DermLite DL1, 3Gen, San Juan Capistrano, CA, USA) for image acquisition and transmission to DERM (Fig. S1). Upon case submission, DERM provided diagnoses and treatment recommendations to GPs, allowing for follow-up questions. Throughout the project, software functionality was expanded, based on practicality and security, e.g. including dictation and advanced search functions.

### Study design and televisit process

Patients with unclear skin lesions were informed by their GPs about the possibility of a televisit and in case of written consent the GP recorded the case including medical history, medication, and clinical and dermoscopic images (Fig. 1). GPs also recorded the time spent (in minutes) and rated usability with grades from 1 (very good) to 5 (insufficient). DERM were notified via SMS or email and provided free-text diagnosis and treatment recommendations, preferably within 2 working days. Additionally, the course of action, was systematically classified into six categories, analogous to triage (Fig. 3): (1) no therapy indicated, (2) therapy by GP, (3) routine DERM appointment in <3 months, (4) urgent DERM appointment in <3 days, (5) referral to hospital, or (6) clarifications/others.

**Fig. 1:**
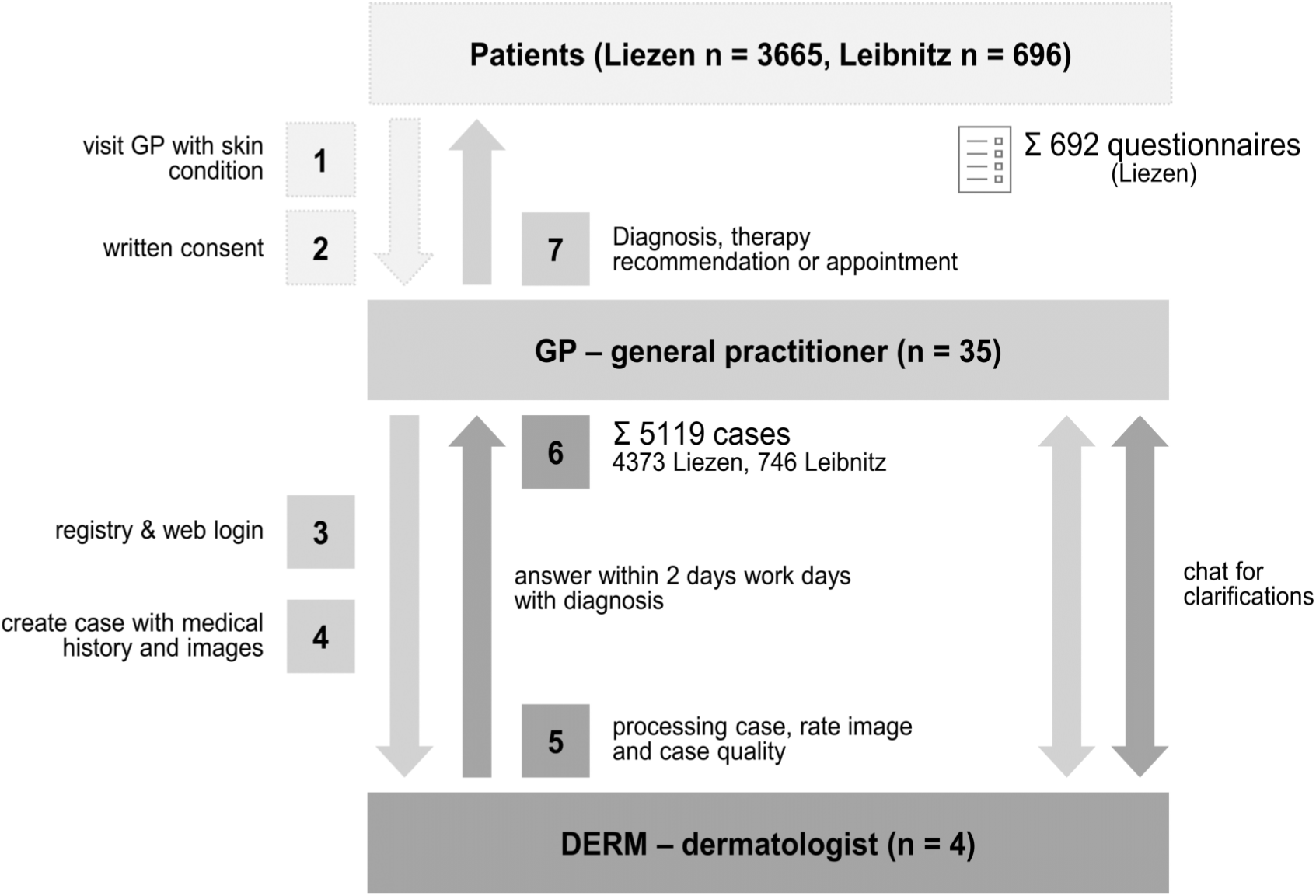
Schematic overview of the teledermatological workflow, detailing the number of cases, GPs, DERM and returned patient questionnaires.

Category (6) primarily involved **DERM** requesting a discussion or additional images, if absent or of insufficient quality. For categories (3-5), **DERM** generally included appointment suggestions. The therapeutic recommendation (2) detailed dosage and duration as free text. Follow-up questions were clarified via the chat function. Upon case closure DERM recorded their time spent (in minutes) and rated the GPs’ image and case quality with grades from 1 to 5.

Patient acceptance of teledermatological visits in Liezen was assessed with questionnaires (Appendix 1) after treatment. GPs from both regions (Liezen, Leibnitz) also received questionnaires, differing for those already participating in teledermatology and non-participants (Appendix 1).

For statistical analysis, all diagnoses were additionally categorized, following a dermatology and venereology textbook^21^, into five main groups (Fig. 3): Inflammatory skin diseases (infections, external factors or unknown cause); cutaneous neoplasm (benign or malign); localized skin conditions; vascular diseases; others.

### Project cost

The Styrian Health Fund covered the costs for IT infrastructure, software, hardware, project coordination, training, ongoing support for participants, scientific guidance, data analysis, and reporting. The medical services for the 5119 cases were reimbursed by the Austrian Health Insurance Funds. Billing was permitted once per patient and per quarter even for multiple televisits, regardless of the additional time and effort required per case.

### Statistics

For statistical analysis, all cases from the Liezen/Leibnitz regions between January 1, 2020, and December 31, 2024, were exported. Technical tests (41), erroneous requests (7), and requests with missing (8) or insufficient (3) images were excluded, resulting in 5119 cases from 4361 patients used for analysis. The age distribution of the general population in 2022 was sourced from Statistik Austria (data.statistik.gv.at/2022_Bev_Alter_Geschl_Bundesl.ods, accessed 11.12.24) and used for comparison.

Statistical analysis utilized R (v 4.4.0) ^22^ and TIBCO Spotfire (v14.4.0, TIBCO, Palo Alto, CA). The age distribution was assessed with Pearson’s Chi-squared test *stats::chisq.test()* with subsequent post-hoc pairwise comparisons *epitools::oddsratio()* and Benjamini-Hochberg (BH) multiple test correction *stats::p.adjust()* analysiert wurden. All data is provided in Appendix 2 with random, pseudonymized IDs not known to anyone outside the research team.

## Results

### Project progression

The “Teledermatology in Styria” project launched on January 1, 2020, in Styria’s Liezen district with 15 GPs and 2 DERM (Fig. 1, Fig. S1). The project was subsequently expanded for additional GPs in Liezen and the district Leibnitz to a total of 35 GPs und 4 DERM (Fig. 2). After 5 years 5119 completed cases were evaluated. Of the 35 GPs, 2 withdrew due to retirement, while 6 declined to continue teledermatology after a test phase and vacancies were reoccupied.

**Fig. 2:**
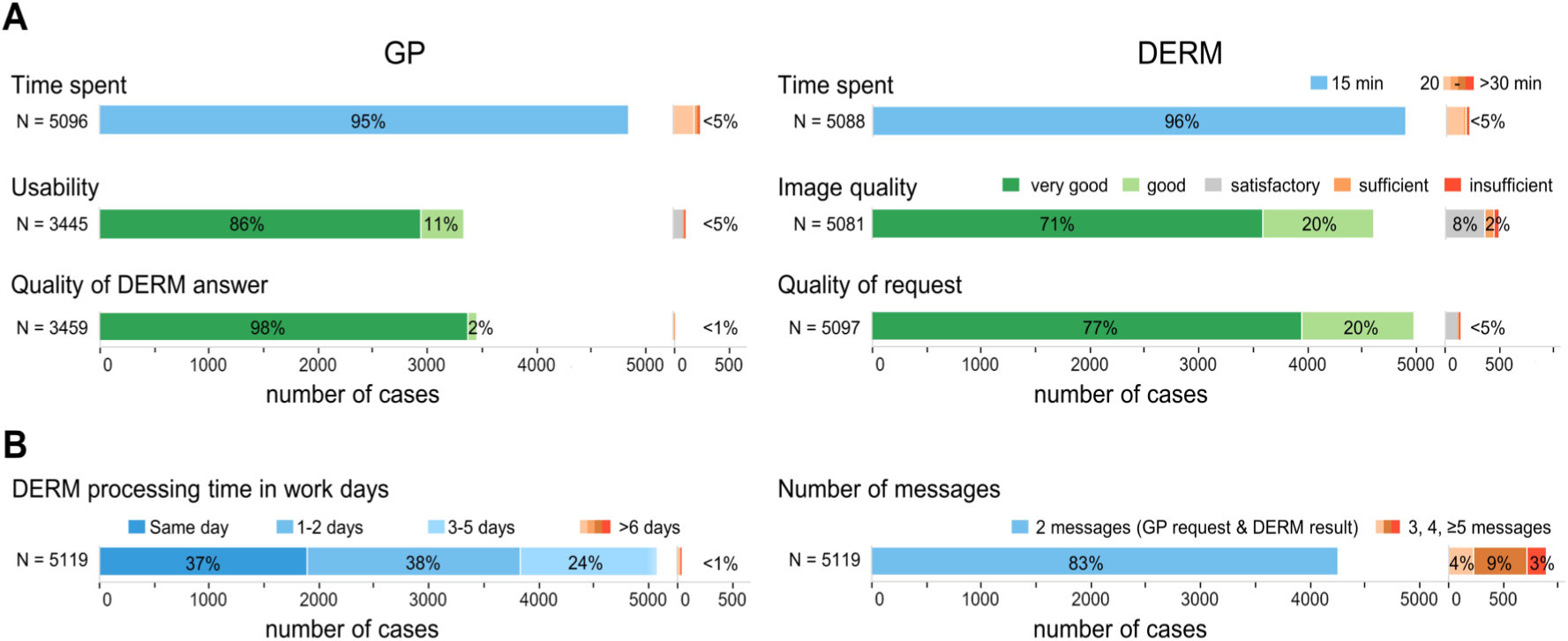
Quality, time spent, and satisfaction of GPs and DERM with the teledermatological system. **(B)** Summary of case-specific evaluations as percentages of all completed cases. (B) Summary of DERM processing time in working days (excluding weekends and holidays) and the number of messages exchanged per case. No distinction was made between medical inquiries and thank-you messages.

### Evaluation of the teledermatological system from the perspective of GPs and DERM

GPs and DERM completed 95% and 96% of case submissions and responses within 15 minutes, respectively (Fig. 2). In 86% of cases GPs rated the usability as very good (user interface in Fig. S1) and in 98% of cases GPs rated the DERM responses as very good (Fig. 2). DERM rated 71% of images and 77% of case requests as very good. Three-quarters of DERM responses were provided within 2 working days, and 17% utilized the chat function for follow-up questions (Fig. 2).

The annual case frequency per GP remained largely stable, without relevant reduction or increase. While only 38% of GPs were female, they submitted a similar number of cases (n = 2351) as male GPs (n = 2465).

### Demographics, diagnostic categories, and triage

The gender and age distribution of the 4361 patients from the 5119 cases shows that with 56% more female patients participated (Fig. 3). Compared to Austria’s general population (Fig. 3B) there was an overrepresentation of older patients and underrepresentation of younger ones. Both districts had nearly identical demographic cures, except for relatively more children being included in Liezen. With the continued availability of the teledermatological system, up to one-third of patients returned over the years (Fig. 3C), but 69% presented with a different skin condition.

**Fig. 3:**
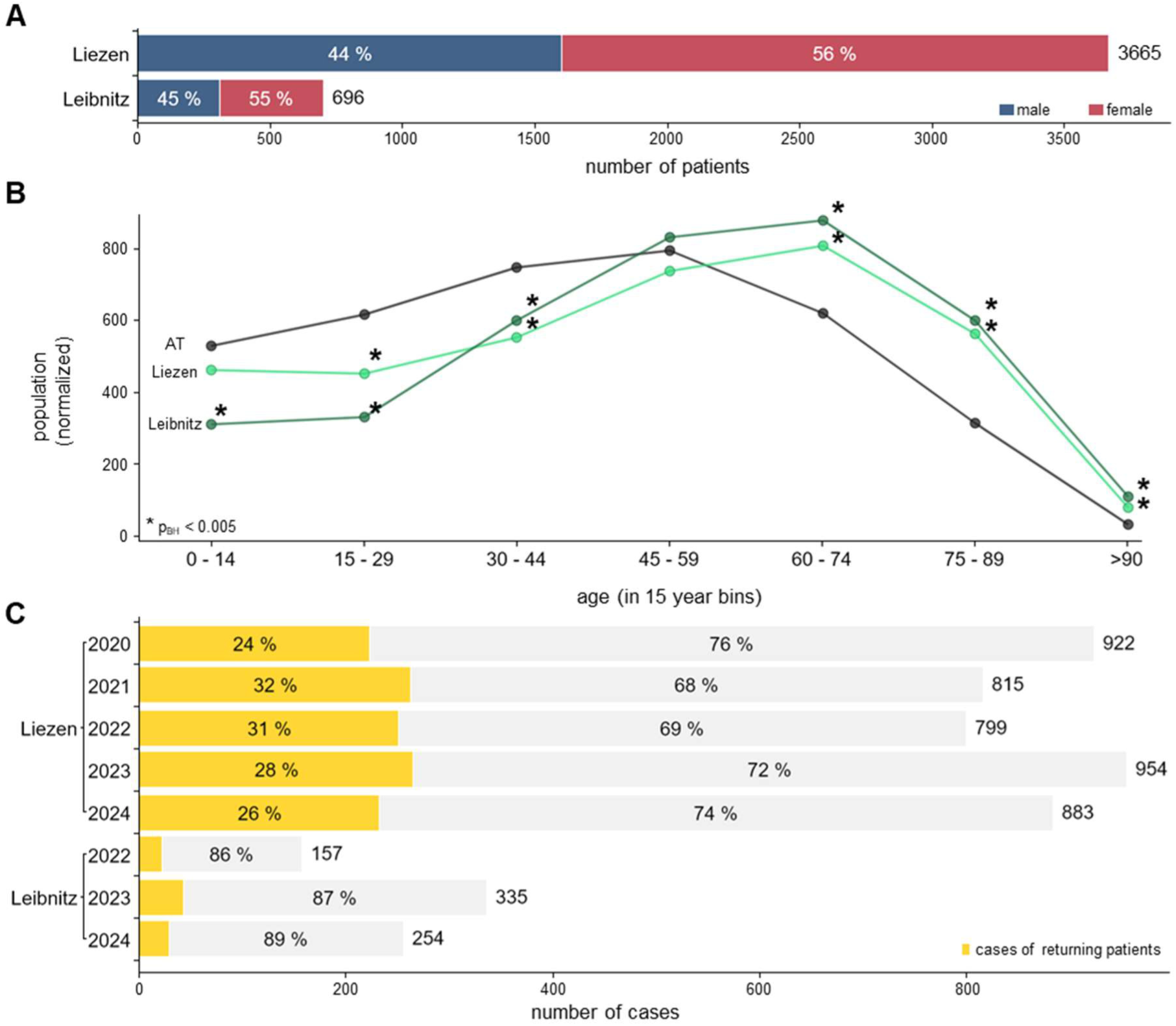
Overview of distributions of patients and cases. **(A)** Distribution of patients by gender and region. **(B)** Age distribution of patients compared to the Austrian general population (AT). For better comparability, AT and Leibnitz were normalized to the total count in Liezen. The two-sided Chi-square test for Liezen vs. AT and Leibnitz vs. AT was significant (p < 0.001). The results of the post-hoc pairwise comparisons within the age group, using 45-59 as the reference group and Benjamini-Hochberg (BH) multiple test correction, are indicated by an asterisk (p_BH_ < 0.005). **(C)** Distribution of cases per region and year, with the number of returning patients highlighted in yellow.

In 23% of all 5119 cases GPs did not provide a suspected diagnosis, in 25% the GPs diagnosis matched the DERM diagnosis and in 51% diagnosis was changed by DERM (Fig. 4A). In 19% of cases no therapy was needed, while in 61% therapy by GP was possible, 12% needed a routine dermatological appointment, 3% an urgent appointment, and 2% a hospital appointment. Consistent with an efficient triage paradigm, 80% of all cases were successfully concluded via teledermatology, with only 17% necessitating referral to DERM or a hospital (Fig. 4B).

**Fig. 4:**
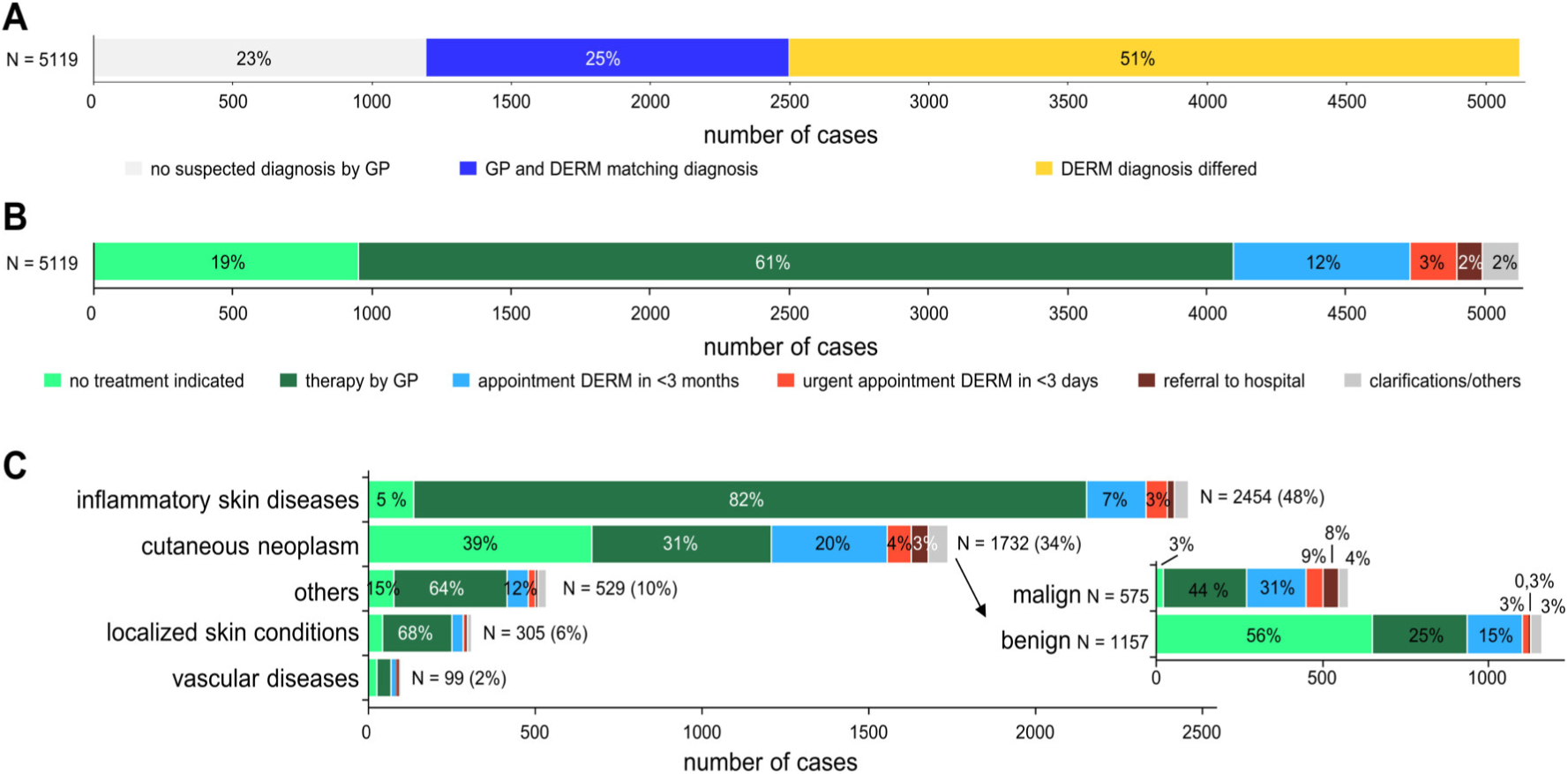
Televisits enable efficient triage. **(A)** Comparison of the suspected diagnoses by GPs with DERM diagnoses. **(B)** Overview of subsequent case management based on DERM diagnosis. **(C)** Different diagnostic groups differ in their percentages of necessary therapy or referrals.

Categorization of the diagnosis revealed that 34% (n = 1732) of cases were neoplasms, of which 67% were benign (n = 1157, Fig. 4C). Malignant neoplasms constituted 33% (n = 575), with nearly all receiving treatment: 44% by GP and 48% by DERM. Only 8% declined treatment for various reasons such as palliative care. The vast majority of cases (n = 3387) involved conditions from the broad dermato-venereological spectrum. Most of these also required treatment, though an additional examination by DERM was rarely necessary (Fig. 4C).

Neither the gender of the patients nor the gender of the GP led to any relevant differences in subsequent therapy or referral frequency (Fig. S2).

Remarkable differences were observed in the urgency of further treatment between the various specific diagnoses, with malignant tumours presenting highest urgency. In contrast, inflammatory skin diseases, including inflammatory tumours (dermatofibromas, keloids, viral warts, abscesses, etc.), were almost entirely treatable by GPs (Fig. 5).

**Fig. 5:**
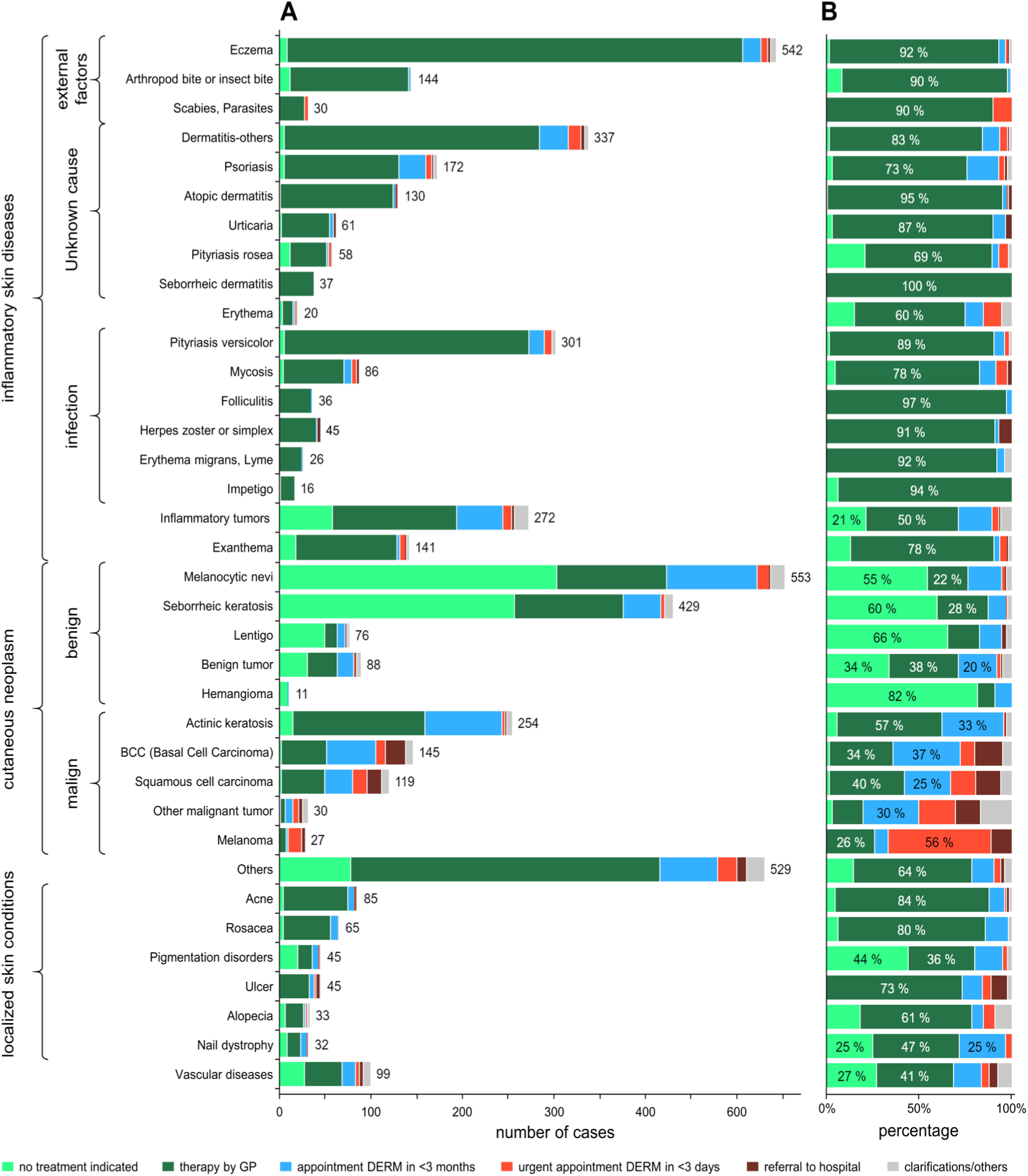
Urgency varies significantly by specific diagnoses. **(A)** Case numbers display the varying prevalence of specific diagnoses. **(B)** The percentage distribution of subsequent therapy or referrals per diagnosis highlight the large differences in urgency. Especially inflammatory diseases have a high demand for therapy, which is mostly possible by GPs.

### Evaluation by questionnaires

A total of 3665 questionnaires were distributed to patients in Liezen, of which 692 (18.8%) were returned after completed teledermatological treatment. The demographic distribution shows good representation across all age groups, genders, and educational levels (Fig. 6). More responses were received from female than male patients (48% to 38%). Overall, 95% of patients were satisfied with the teledermatology service. After treatment, satisfaction with the results was 87% (Fig. 6B), with only 1% responding no and another 12% providing no answer, partly because their treatment had not yet begun.

**Fig. 6:**
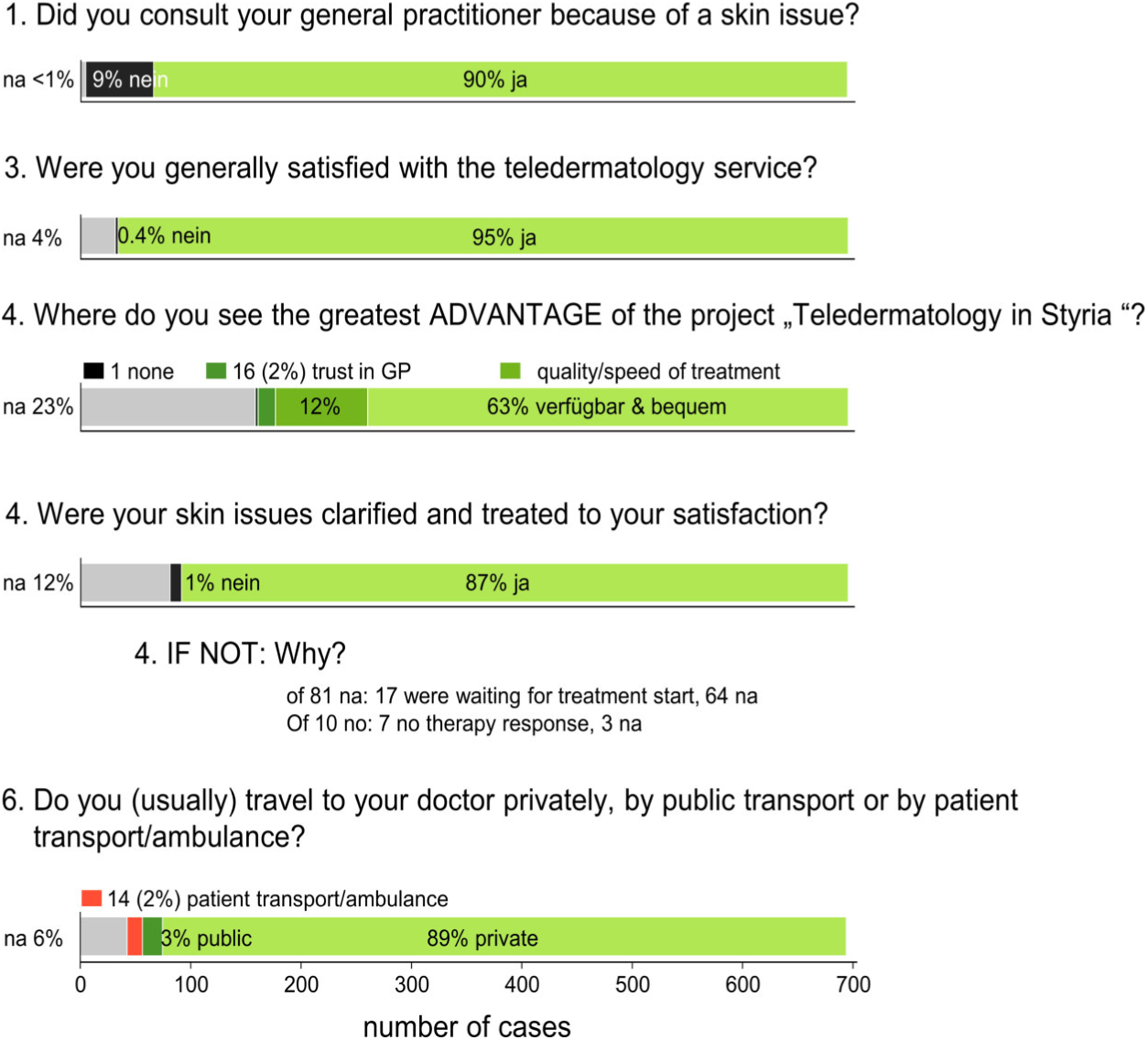
Patient satisfaction from questionnaires. Results show patient behaviour and satisfaction based on answers within the returned questionnaires. na – no answer provided

The free text field „greatest advantage” elicited responses from 77% of patients. Easy access and convenience were the most frequently mentioned benefits, alongside trust in the GP and the quality/speed of treatment (Fig. 6). Disadvantages were not reported by 96% of patients, while 3% criticized the lack of personal contact with DERM (Fig. S3). While most patients travelled privately (89%) or via public transport (3%), 2.3% (14 patients) required an ambulance or patient transport (Fig. 6). Detailed information from the questionnaires, such as distances, time savings, or the method of information dissemination to patients, is summarized in Fig. S3 and is detailed in Appendix 2. A total of 34 patients made the effort to share personal comments, all of which were positive and expressed gratitude, wishes for service expansion, or praise.

GPs from the districts Liezen und Leibnitz received questionnaires, separated by GPs without and GPs with teledermatological service (questionnaires Appendix 1, data Appendix 2). From the GPs with teledermatological service 51% (n = 16) of the questionnaires were returned. The average reported satisfaction was 1.25 (very good) with only one mark 3 (satisfactory). All GPs saw advantages, primarily emphasizing speed, time savings, short processing times, low-threshold accessibility to diagnosis and treatment guidance as well as a learning effect. Almost all GPs (15 of 16) strongly endorsed the system and would recommend it to their colleagues. The chat function was used by 7 GPs for clarifications, reporting complete satisfaction with the responses received.

Questionnaires were also send to 86 GPs in Liezen and Leibnitz without access to the teledermatological system, of which 23% (n = 20) were returned. Just 4 GPs were unaware of the ongoing teledermatological project, while 15 expressed their desire to participate in the project.

GPs without teledermatology anticipated advantages consistent with those reported by already participating GPs, and additionally highlighted the connection to the hospital. However, GPs without teledermatology listed more disadvantages, such as the lack of direct patient contact, additional administrative work, limited diagnostic capabilities from poor image quality, and increased workload for AMs.

## Discussion

Our aim was to develop a pragmatic solution to overcome rural healthcare shortages, caused by the high prevalence of dermatological diseases^23–26^ and the, since decades, steadily increasing dermatological care burden^20^. The current GP-to-DERM referral system creates prolonged waiting times, leading to severe patient morbidity and a higher economic burden.

A total of 35 GPS from districts Liezen and Leibnitz were digitally connected with 4 DERM via a SF-technology. Thus all communication within this project was exclusively between medical practitioners. The project aimed both to improve interdisciplinary cooperation and to ensure faster access to dermatological care by triaging patients by disease severity. This teledermatologically enhanced communication, was designed to strengthen existing networks, maintain patient flows, save on both time and travel, and enhance dermatological care.

Therefore, patients had no in-person contact to DERM, but rather their case was sent for DERM assessment, including images and medical history, if deemed necessary by the GP. This effectively replaced the traditional referral process to a DERM with a digital referral. After the case was answered the GP handled diagnosis consultations and therapy administration, saving time and avoiding “being left on your own” of patients. This lack of support is the major issue with internet-based self-diagnosis or medical advice where a plethora of both trustworthy and dubious medical services are offered side-by-side. Intriguingly, even artificial intelligence (AI)-supported tools, often considered diagnostically reliable, frequently caused anxiety rather than reassuring patients ^27,28^. Studies found that patients forget 40-80% of medical discussions shortly after leaving, which highlights the GPs’ key role in providing primary medical care, documenting relevant information, and reiterating details as needed.

Through this project, we have successfully established a teledermatological service primarily for outpatient care, a service that not only integrates with but also improves existing healthcare structures. The collaboration among policy makers, health insurance funds, medical associations, and medical practitioners from both outpatient and hospital is unique. Also distinctive is the five-year duration of the study with comprehensive data, featuring per case resolution for 5119 cases and a high number of returned patient questionnaires. The level of detail per case, e.g. categorizing the course of action – ranging from no therapy to urgent referrals including appointments – and reporting time spent as well as quality grading is distinguishing compared to other large studies^29–31^.

Over 90% of both GPs and DERMs rated user-friendliness as very good or good. A remarkable efficiency was observed, as over 95% of cases were submitted (GP) or answered (DERM) within 15 minutes.

Most surprisingly only 17% of all cases require an additional in-person dermatological consultation (Fig. 4). In 61% of cases therapy was led by GP according to DERM recommendations, the most common course of action in inflammatory skin diseases. No therapy was indicated in 19% of cases, mostly for benign tumours. These distribution was mostly constant over the project duration.

The diagnostic group breakdown revealed the unexpectedly low proportion of one-third being tumours, of which only one-third were malign. Within those malignant tumours the actinic keratosis was the largest subgroup and was largely amenable to therapy by GPs. Dermatoses, predominantly inflammatory, were the largest diagnostic group, required mostly therapy, which was mostly managed by GPs.

Certain dermatoses present initially with minimal discomfort often leading to late diagnosis and consequential damage. Examples include early-stage malignant tumors and infectious conditions such as Borreliosis. *Erythema chronicum migrans* was identified in 26 cases and is considered cured after a two-week antibiotic therapy. Conversely, untreated Borreliosis can affect various organ systems ending in invalidity, thus being a recognized occupational disease in Austria. This year, the definition of recognized occupational dermatoses was expanded to include tumours caused by chronic UV exposure. Early diagnosis and intervention for these conditions substantially reduce health care and public costs. Furthermore teledermatology saves costs in patient transport, with 2.3% of patients reportedly relying on patient transports or ambulances for their appointments (Fig. S3).

Notably fewer cases were submitted in Leibnitz compared to Liezen relative to their population. Topographically, Leibnitz is closer to areas with better access to dermatological care. This implies that this form of teledermatology is applied only when genuinely necessary, primarily in dermatologically underserved, thinly populated regions requiring long travel.

In previous literature the digital inequality is described as limitation of telemedicine. This socially and geographically induced inequality restricts digital access to specific demographic groups^32^. However, this inequality is resolved by the direct digital connection of GPs with DERM. Crucially, the inherent challenges of SF-technology^33^ related to collection of medical history, in-person contact and direct diagnosis or therapy consultations were effectively mitigated by involving knowledgeable GPs. The teledermatological involvement to the Universities’ Department of Dermatology in Graz ensured that every case was ultimately resolved. In ambiguous cases, the diagnosis and treatment recommendations were supported by a wider medical consensus, ensuring no one – neither patient, AM, nor DERM – were left alone with a problem. This is in contrast to Otten’s study in 2023^34^ on a German teledermatological service, which was able to clarify 70% of the cases but satisfactory resolution was only achieved for 53%^35^. In our project, 95% of surveyed patients positively viewed the teledermatological service, and 87% were satisfied with the management of their skin problem.

While the GPs’ workload increase is undisputed, it is offset by the efficiency gains from the triage effect. DERM can integrate the teledermatological work quite well into their clinical routine with proper scheduling. Unlike direct in-person visit within an often hectic clinical setting, SF-teledermatology enables focused and uninterrupted work. The administrative workload increases for GPs, but decreases for DERM compared to in-person consultations. However, crucially this is outweighed by obtaining considerably earlier a specialists diagnosis and intervention. In case of insufficient image quality, it is the DERM’s responsibility to request better images or to refer the patient for an appointment. The responsibility for the correct implementation of DERM recommendations, lies with the GP. Thus for both parties, an appropriate extension of liability insurance is advisable. Although, we have not recorded a single case of liability in five years.

These consistently positive experiences served as a strong motivator for the authors, all GPs, and DERMs throughout the project’s planning and implementation, simultaneously explaining the project’s success: a healthcare gap was closed, regional structures were utilized, and costs were reimbursed. Ultimately, the success is rooted in the effective, constructive planning and the shared determination of all stakeholders to succeed.

The routine reimbursement of the medical services by health insurances is unique, but does not reflect the actual workload. The paid reimbursement falls somewhat short of the value reported in a comparable 2018 Australian study^36^, which only assessed tumours. Treatment of complex dermatoses by GPs, as it was routinely performed in our project, was not part of the Australian study. This emphasizes this project’s worth: A legally compliant, secure teledermatological tool empowers GPs and DERM to efficiently process and resolve every case. This direct physician-to-physician communication of equals solves real-world problems and has now truly become integrated into daily clinical life.

## Limitations

This study has a non-randomized design, lacking comparable reference regions. Consequently, data on the true dermatological referral rates was unavailable. Also, the observation period spanned the COVID-19 pandemic and subsequent years, which, due to the continuous scaling of the project, meant the pandemic’s influence could not be distinctly isolated.

## Data Availability

All data is provided in Appendix 2.

## Acknowledgements

The authors sincerely thank all patients and general practitioners (Heidelinde Altenaichinger, Anneliese Auer, Wolfgang Auer, Dominik Augustin, Philipp Brodatsch, Claudia Burgstaller, Astrid Doppler, Jakob Dorner, Pia Edlinger, Walter Gsöllpointner, Daniela Habersatter-Theil, Markus Hamp, Martin Hasibeter, Johann Holler, Norbert Holzmüller, Renate Jocham, Thomas Kappaun, Klaus Karrer, Oliver Lammel, Manfred Manninger, Doris Nestl-Treiber, Magdalena Putz, Martina Rauscher, Richard Rezar, Josef Rampler, Alexander Schwarz, Peter Sigmund, Christian Soral, Robert Sponner, Alexander Stradner, Maria Thier, Sabine Trummer-Grün, Renate Wiesler, Markus Wippel, Thomas Zorn, Gudrun Zweiker) and all dermatologists (Urban Cerpes, Peter Kahofer, Thomas Kainz, Manfred Tritscher) for their long-term dedication, invaluable feedback, and data donations. We wish to express our deep appreciation to Franz Schuller and Horst Stuhlpfarrer for their many productive discussions, essential technical support, and outstanding work.

## Declaration of generative AI and AI-assisted technologies

During the preparation of this manuscript the authors used large language models (LLMs, e.g. Gemini, DeepL) to aid translation and improve readability of some text. The authors reviewed and edited afterwards meticulously the text and take full responsibility for the content of the publication.

## Appendix 1

### Security

The application provided by E-derm-consult is general data protection regulations (GDPR) compliant with yearly exerternal audits and the company is ISO 9001-2008 and ISO 13485:2003 certified. Both, GPs and DERM can only be registered by the technical administrator and both always have to login via username/password with two-factor authentication from any internet-capable device.

Data transfer is encrypted (HTTPS connection), and all patient-related data and images are stored exclusively on E-derm-consult’s Austrian servers, not on the iPads. Data backups are performed revolving daily, weekly, monthly, and annually on a separate server. The databases are fundamentally encrypted at the data file level, and exports for scientific analyses can only be released via a key. Passwords are always stored as hashes.

**Fig. S1:**
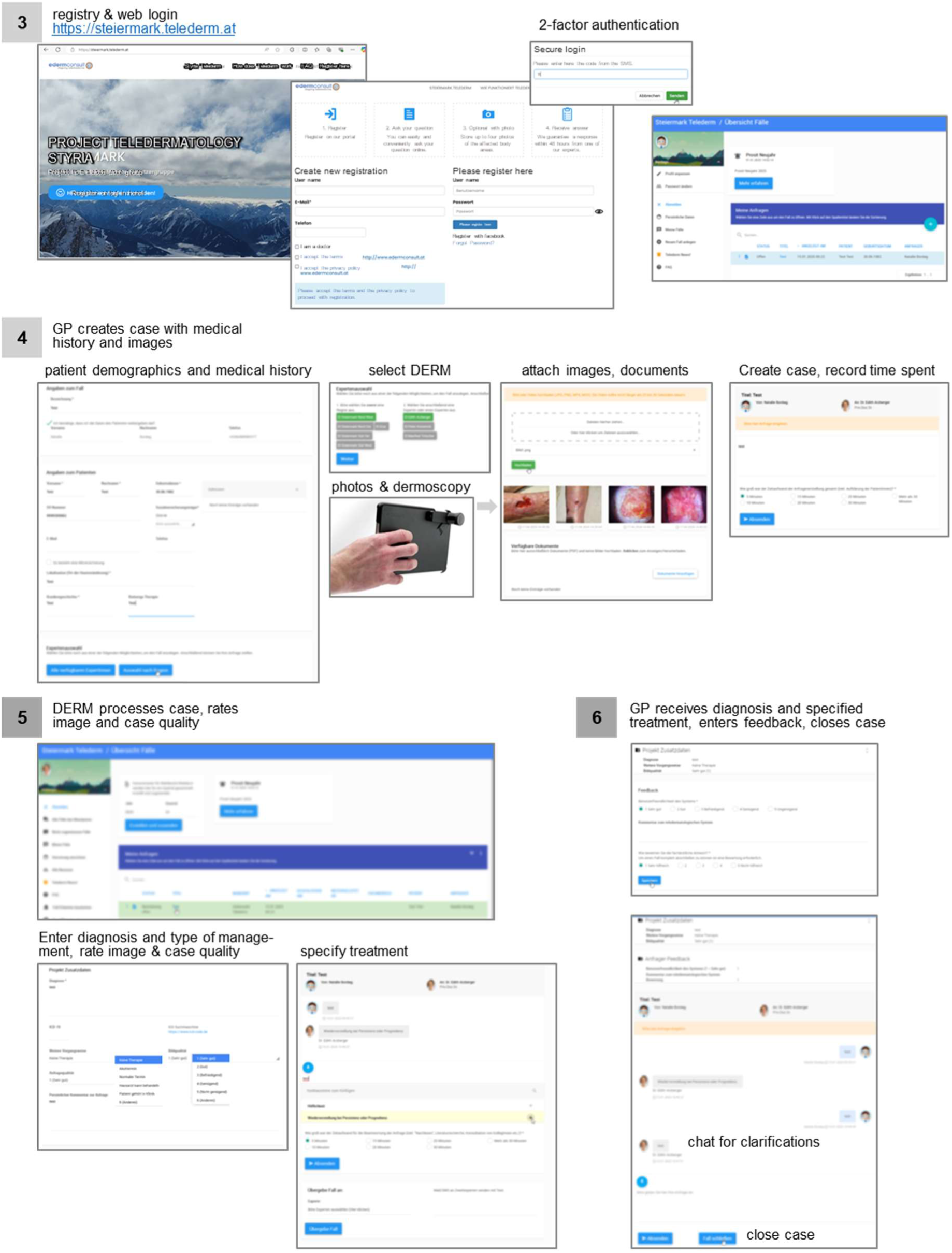
The graphical user interface from the GPs (3, 4, 6) and the DERMs (3, 5) perspective. The numbers also correspond to the steps illustrated in Figure 1.

**Fig. S2:**
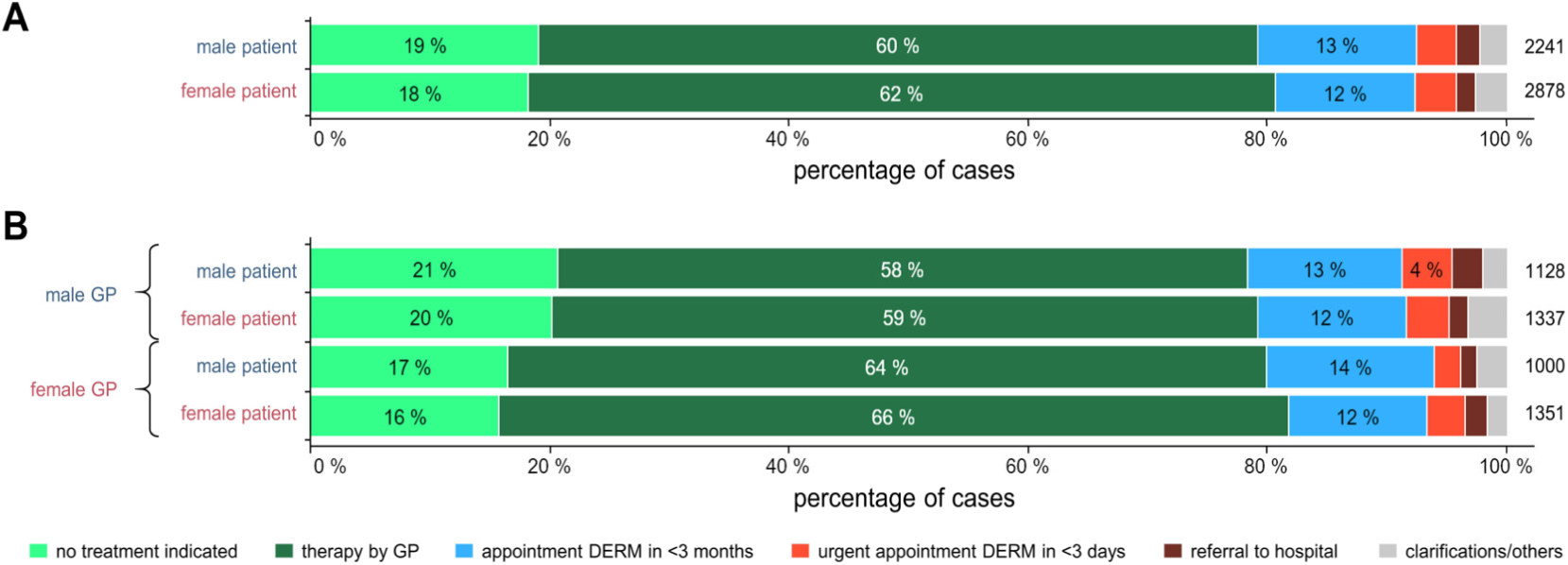
The course of action is independent of the patients’ or GPs’ sex. **(A)** The course of action is independend oft he patients’ sex. **(B)** The course of action is also independent form the GPs sex and their patients’ sex.

**Fig. S3:**
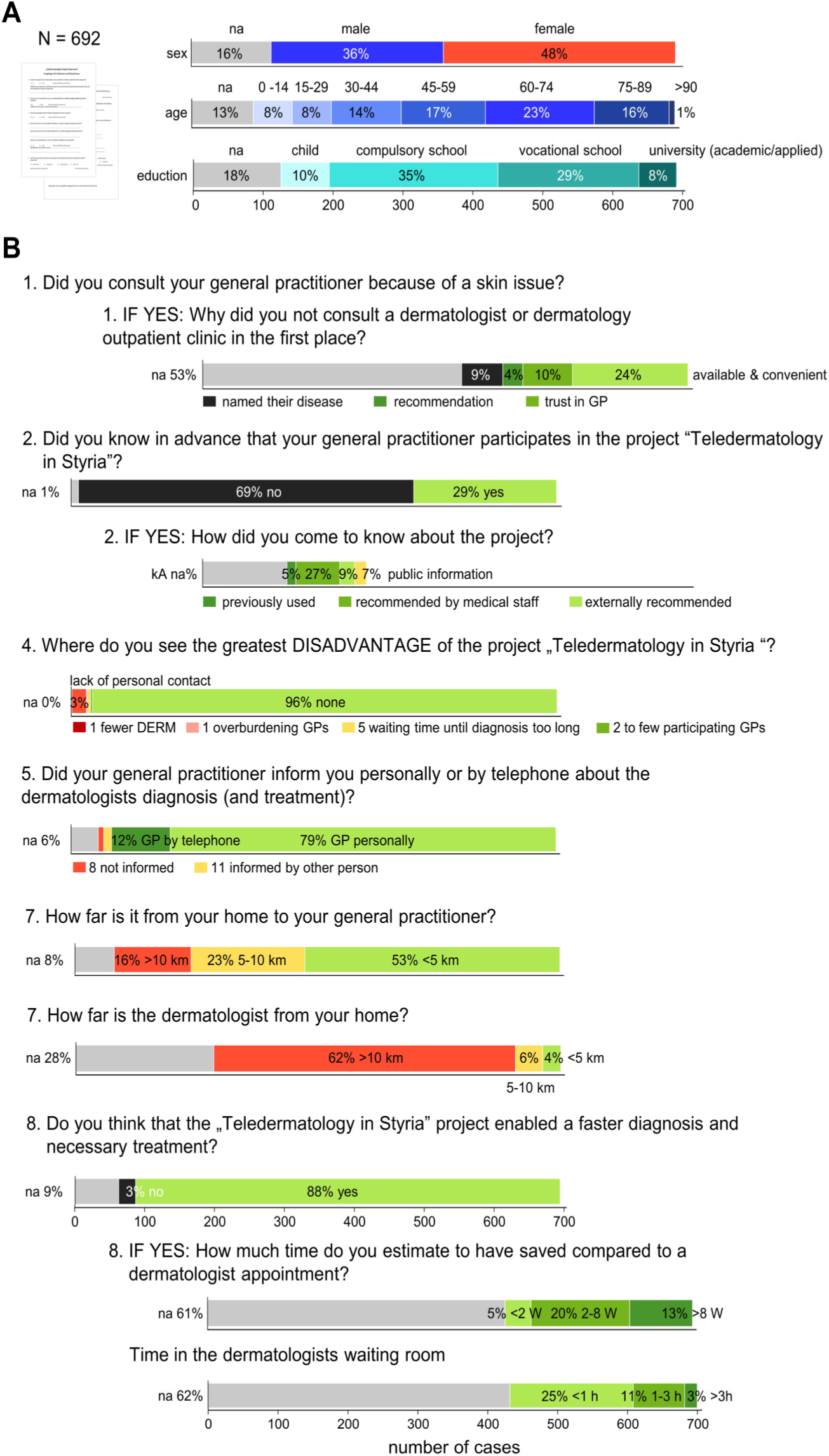
Patient Questionnaire Results. **(A)** Distribution of patient demographic characteristics. **(B)** Results show patient behaviour and satisfaction based on answers within the returned questionnaires. na – no answer provided

### Translated questionaire for patients

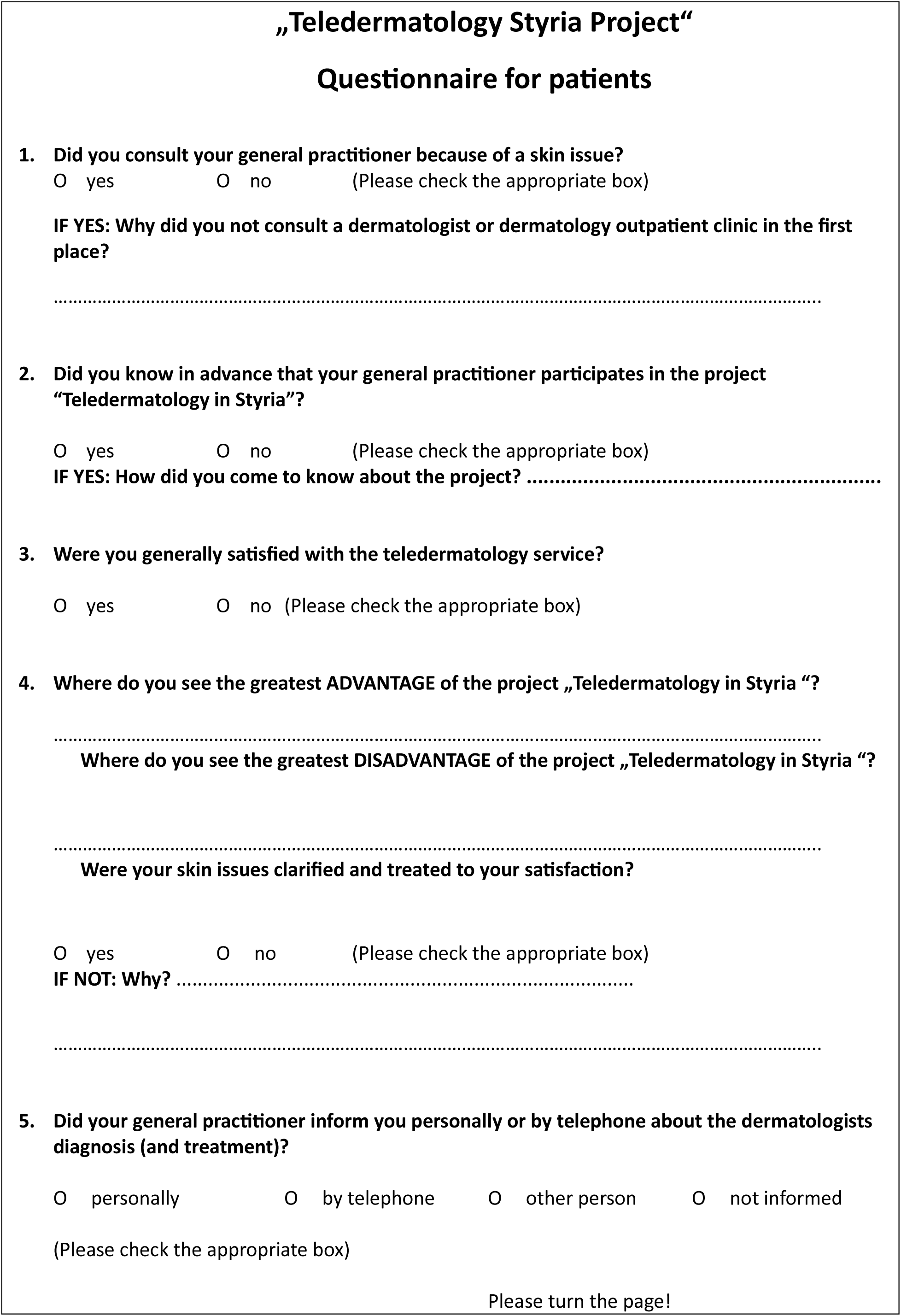
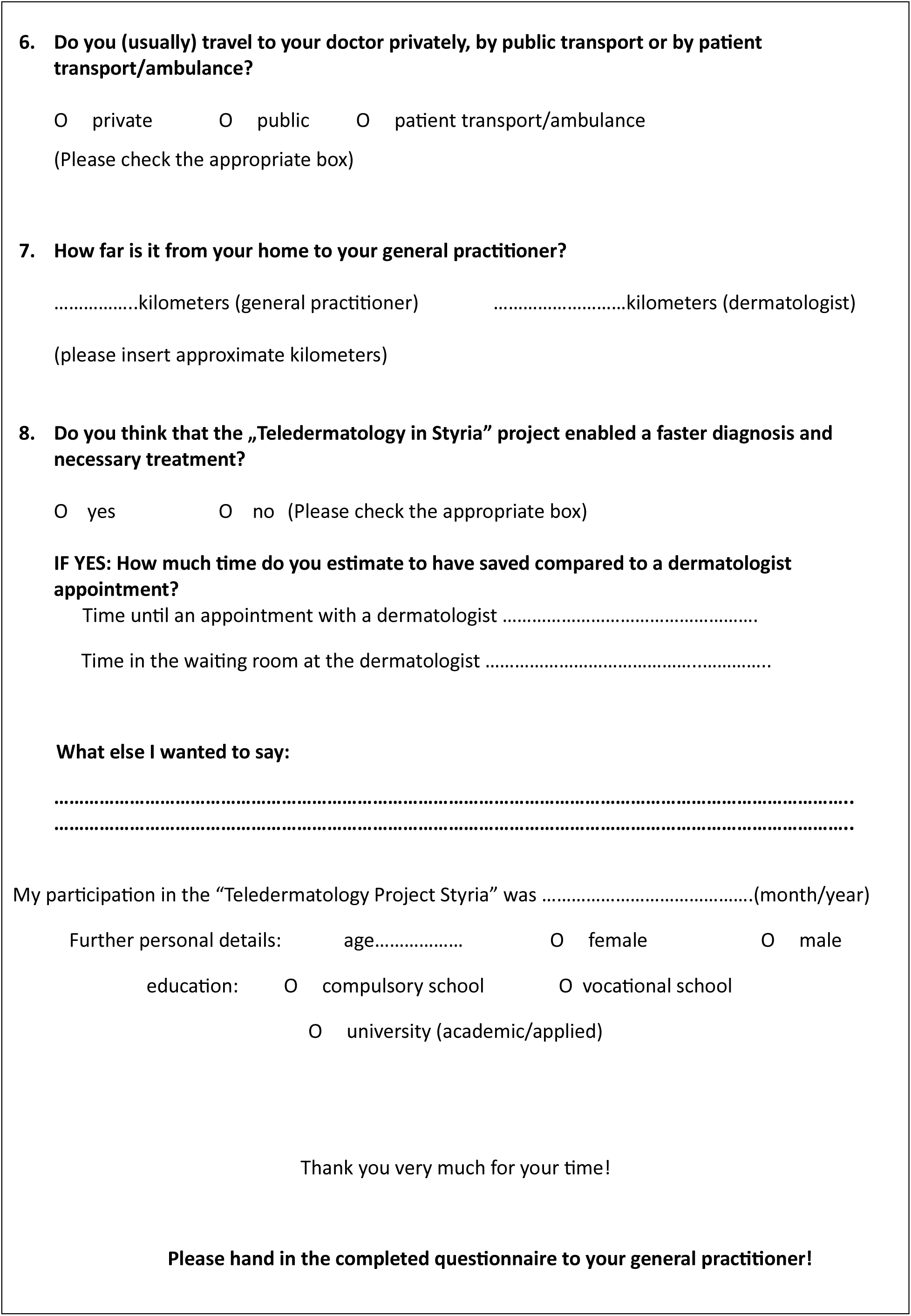

### Translated questionaire for GPs with teledermatological service

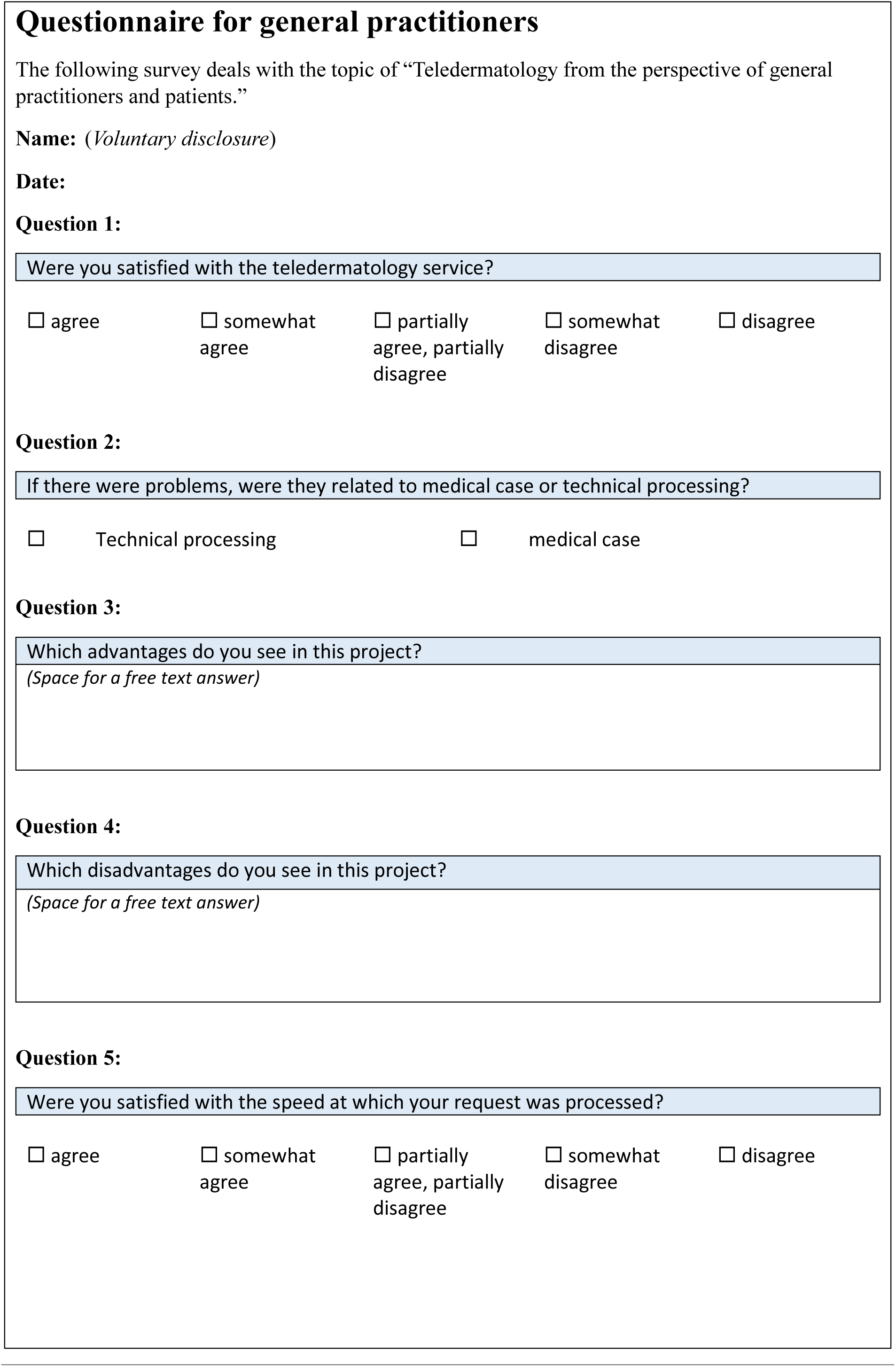
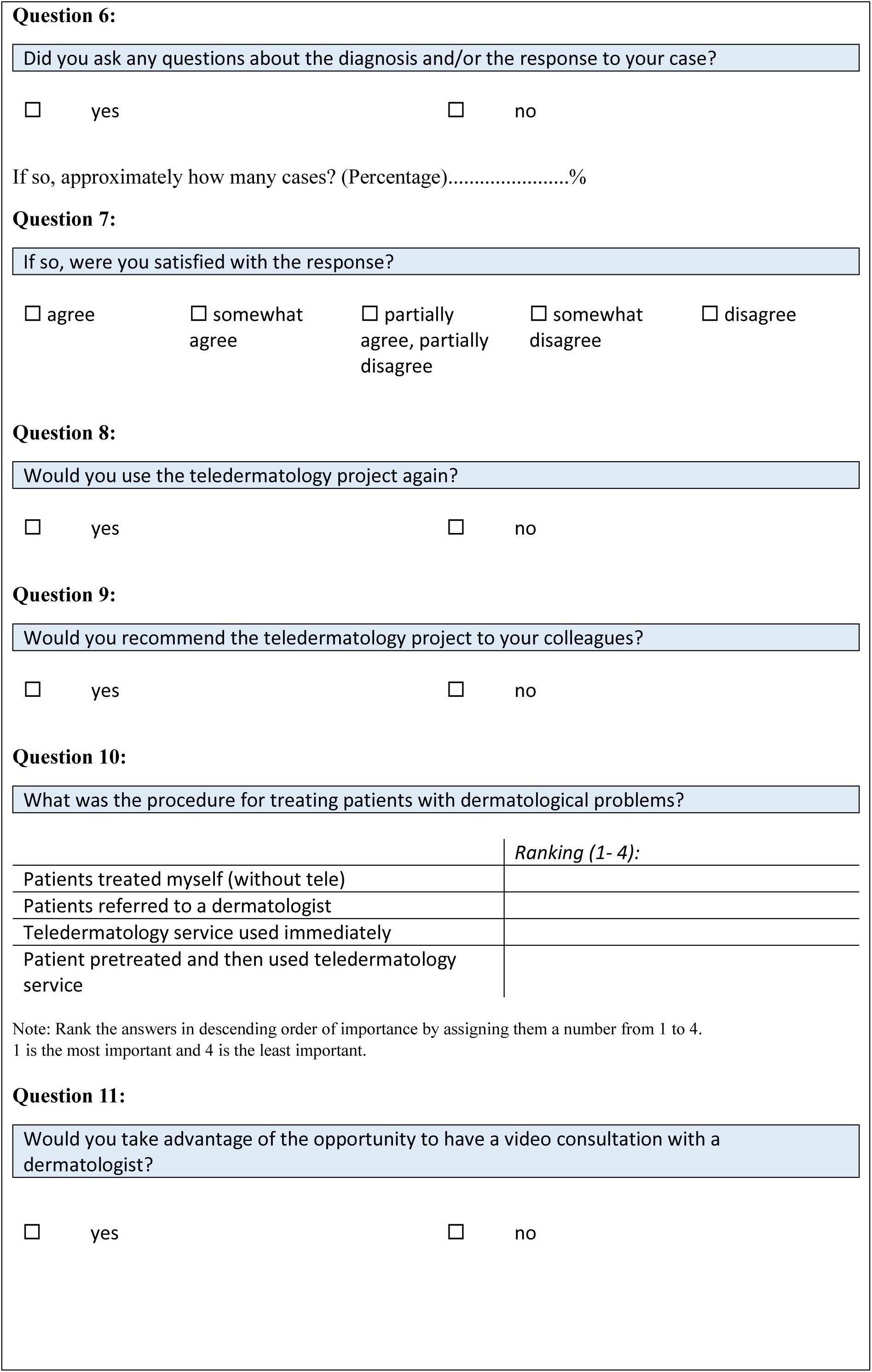
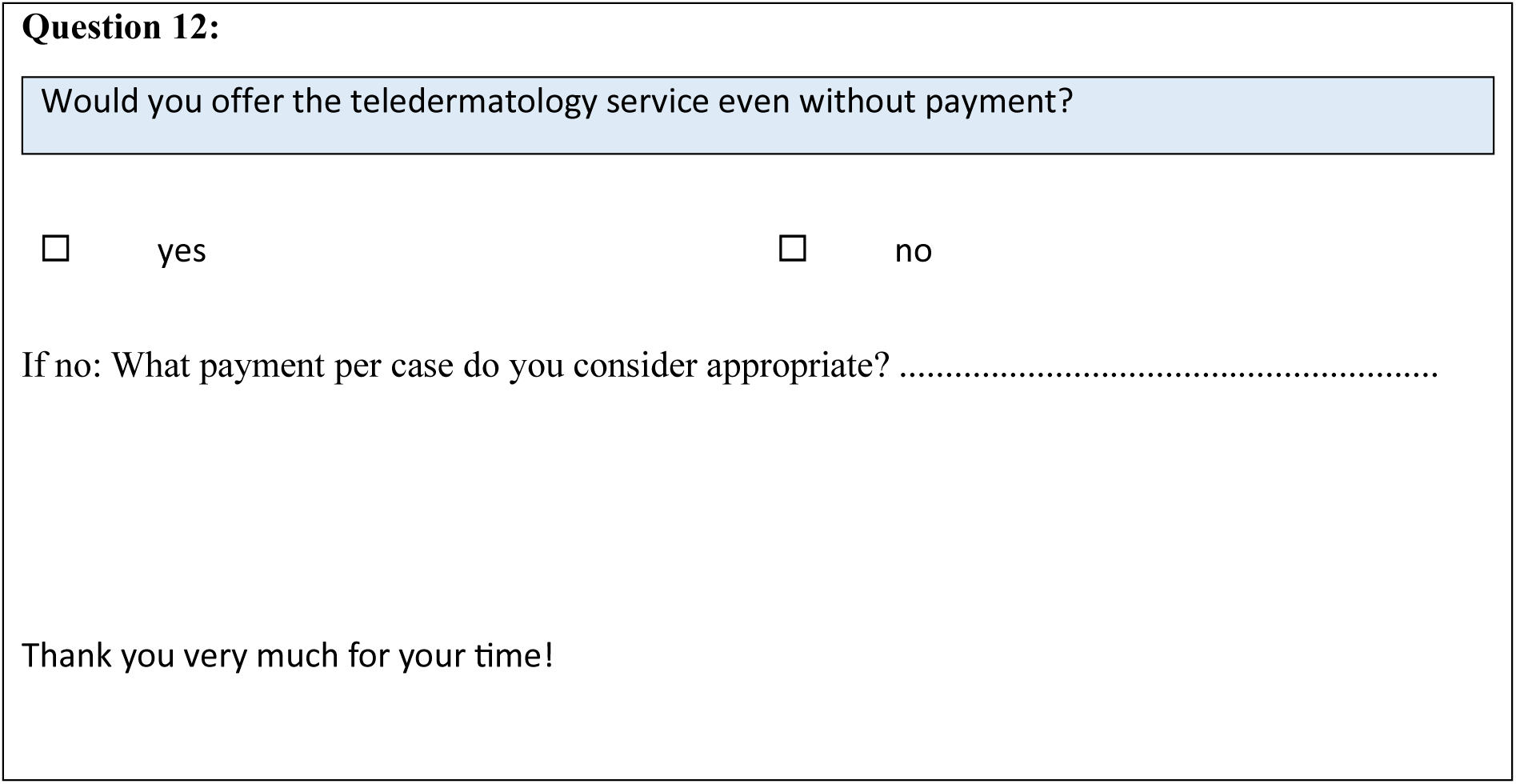

### Translated questionaire for GPs without teledermatological service

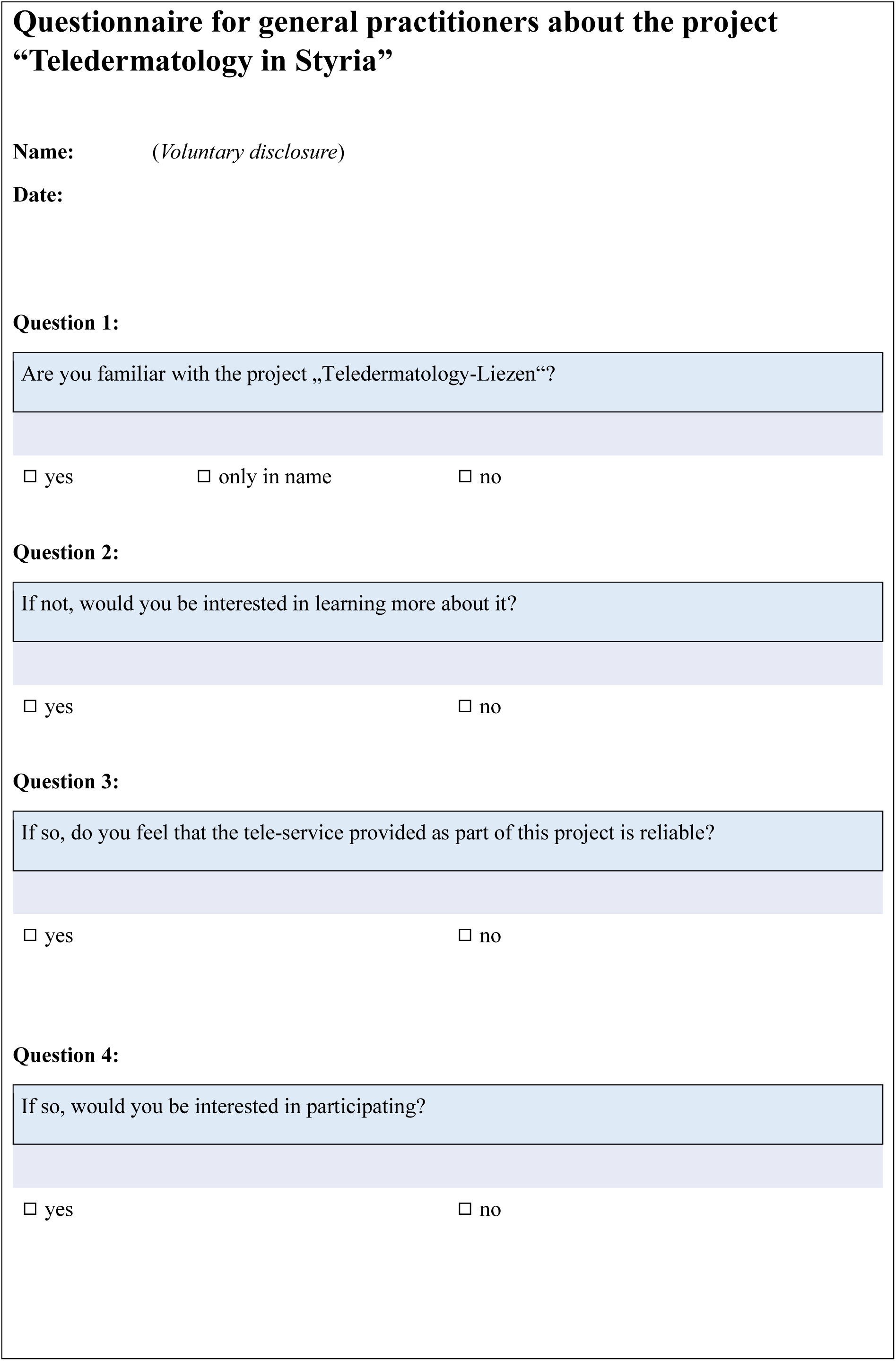
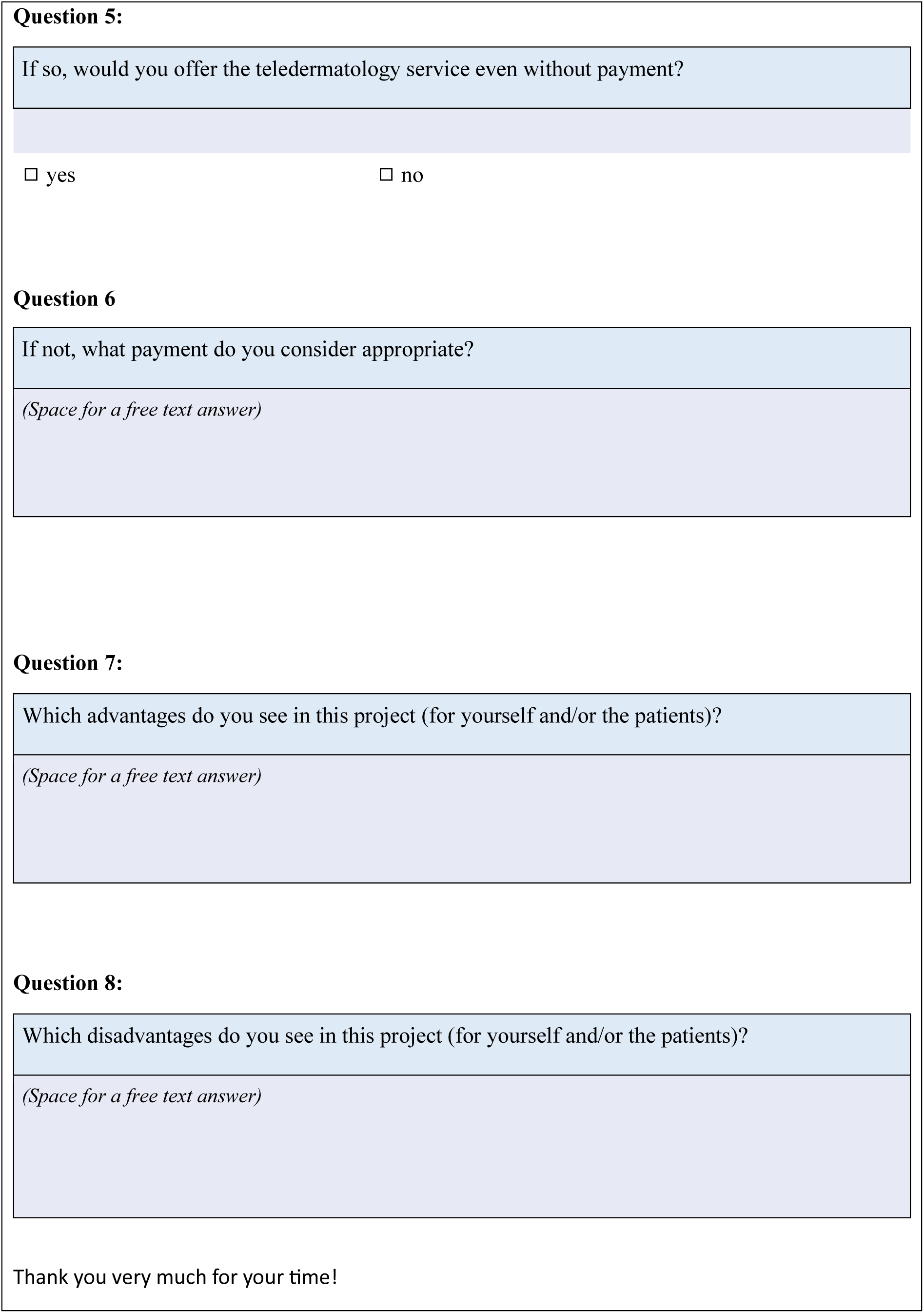

